# Phenome-wide analysis of *APOL1* risk variants reveals associations between one combination of haplotypes and multiple disease phenotypes in addition to chronic kidney disease

**DOI:** 10.1101/2023.02.19.23285950

**Authors:** Walt E. Adamson, Harry Noyes, Paul Johnson, Anneli Cooper, Darren G. Monckton, John Ogunsola, Michael Sullivan, Patrick Mark, Rulan S. Parekh, Annette MacLeod, the TrypanoGEN+ Research Group as members of the H3Africa Consortium

## Abstract

**Background:** Infectious diseases are a major driving force of natural selection. One human gene associated with strong evolutionary selection is *APOL1*. Two *APOL1* variants, G1 and G2, emerged in sub-Saharan Africa in the last 10,000 years, possibly due to protection from the fatal African sleeping sickness, analogous to *Plasmodium*-driven selection of the sickle-cell trait. As homozygosity for the HbS allele causes sickle cell anaemia, homozygosity for the *APOL1* G1 and G2 variants has also been associated with chronic kidney disease (CKD) and other kidney-related conditions. What is not known is the extend of non-kidney-related disorders and if there are clusters of diseases associated with individual APOL1 genotypes.

**Methods:** Using principal component analysis, we identified a cohort of 10,179 UK Biobank participants with recent African ancestry. We conducted a phenome-wide association test between all combinations of *APOL1* G1 and G2 genotypes and conditions identified with International Classification of Disease phenotypes using Firth’s bias-reduced logistic regression and a false discovery rate to correct for multiple testing. We further examined associations with chronic kidney disease indicators: estimated glomerular filtration rate (eGFR) and urinary albumin:creatinine (uACR).

**Results:** The phenome-wide screen revealed 74 (mostly deleterious) potential associations with hospitalisation for a range of conditions. G1/G2 compound heterozygotes were specifically associated with hospitalisation in 64 (86.5%) of these conditions, with an over-representation of infectious diseases (including COVID-19) and endocrine, nutritional, and metabolic diseases. The analysis also revealed complexities in the relationship between *APOL1* and CKD that are not evident when the risk variants are grouped together: high uACR was associated specifically with G1 homozygosity; low eGFR with G2 homozygosity and G1/G2 compound heterozygosity; progression to end stage kidney disease was associated with G1/G2 compound heterozygosity.

**Conclusions:** Among 9,594 participants, stratifying individual *APOL1* risk variant genotypes had a differential effect on associations with both kidney and non-kidney phenotypes. The compound heterozygous G1/G2 genotype was distinguished as uniquely deleterious in its association with a range of ICD-10 phenotypes. The epistatic nature of the G1/G2 interaction means that such associations may go undetected in a standard genome-wide association study. These observations have the potential to significantly impact the way that health risks are understood, particularly in populations where *APOL1* G1 and G2 are common such as in sub-Saharan Africa and its diaspora.

## Introduction

People of recent African origin are disproportionately affected by chronic kidney disease (CKD)^1^. This excess risk has been, in part, attributed to the carriage of two independent variants in the apolipoprotein L1 (*APOL1*) gene referred to as G1 and G2^2^. *APOL1* G1 and G2 are common in sub-Saharan Africa and its diaspora, with estimated allele frequencies of 21% and 13% respectively in African Americans^2^, and up to 49% and 21% in sub-Saharan Africa^3^. G1 and G2 are absent or occur at very low frequency in non-African-derived populations, consistent with the hypothesis that these variants arose in West Africa only 10,000 years ago and were subject to selection in that population^2^ prior to spreading to much of sub-Saharan African and its recent diaspora.

G1 (amino acid substitutions S342G and I384M) and G2 (deletion of N388 and Y389) are both found in the same domain at the C-terminus of *APOL1* only 20 bp apart, but are present on separate haplotypes^2^ and are in complete linkage disequilibrium with each other such that haplotypes with both G1 and G2 alleles are either very rare or absent. Haplotypes containing neither G1 nor G2 are termed G0. G1 and G2 are collectively considered to be high-risk variants for deleterious kidney phenotypes: carriage of two alleles is associated with a spectrum of CKD conditions, including focal segmental glomerulosclerosis, hypertension-associated kidney failure, and HIV-associated nephropathy^2,4–6^. Recently, in SARS-CoV-2-infected African Americans, carriage of two high-risk *APOL1* variants has been associated with collapsing glomerulopathy^7^, acute kidney injury (AKI), persistent AKI, and requirement for kidney replacement therapy^8^. Among patients with COVID-19 disease, carriage of two high-risk variants was associated with increased AKI severity and death^9^. Associations between high-risk *APOL1* variants and non-kidney-specific phenotypes have also been described, including a range of cardiovascular outcomes, however, inconsistently^10^. Studies examining associations with high-risk *APOL1* variants focused primarily on African American cohorts: comparable data for other populations with recent African ancestry is limited.

Previously, a phenome-wide study identified conditions associated with the carriage of two-variant *APOL1* genotypes^11^ associated with conditions recorded via International Classification of Diseases, Ninth Revision (ICD-9) and Tenth Revision (ICD-10) codes. Using stringent criteria, they did not detect associations with non-kidney traits, and concluded that *APOL1* likely only operates in kidney-specific pathways.

APOL1 is found only in humans and few higher primates and is expressed as both a secreted high-density-lipoprotein-associating form by the liver, and as intracellular forms by a variety of cell types, including endothelial cells. Prior to its associations with non-communicable diseases, secreted APOL1 had been identified as the trypanolytic protein: a pore-forming serum protein that lyses protozoan trypanosome parasites, protecting humans from infection by many trypanosome species^3^. The two subspecies of *Trypanosoma brucei* that infect humans, *T*.*b. rhodesiense* and *T*.*b. gambiense*, which cause human African trypanosomiasis, have developed specific mechanisms for avoiding lysis by APOL1, either by binding, avoiding, or degrading the lytic protein^12^. Studies examining the effect of the *APOL1* variants in *T*.*b. gambiense* and *T*.*b. rhodesiense* infections have highlighted differences between the genotypes. Resistance to *T*.*b. rhodesiense* infection was associated solely with the G2 variant but not G1, while in *T*.*b. gambiense* infections, carriage of G1 and G2 were associated with decreased and increased risk of severe disease, respectively^3^. Due to the mainly protective association between *APOL1* risk alleles and human African trypanosomiasis it has been proposed that trypanosomes are the selective agent for *APOL1* G1 and G2 alleles in African populations. This is analogous to the classic example of *Plasmodium* selection for the sickle-cell allele of beta globin in individuals with sickle-cell trait^13^.

Association studies examining *APOL1* G1 and G2 have often grouped the two variants together as recessively ‘high-risk’, however for some conditions the different genotypes have distinct phenotypes as described above for African sleeping sickness. Similarly, in a cohort of African American on long-term haemodialysis patients, different *APOL1* genotypes were associated with different rate of progression to haemodialysis^14^. Recently, the G1/G1 genotype (but not G1/G2 or G2/G2) was associated with proteinuria in a cross-sectional population-based cohort in sub-Saharan Africa^15^.

In light of the *APOL1* genotype-specific phenotypes observed for African trypanosomiasis and the spectrum of kidney-related conditions associated with APOL1 we hypothesised that different combinations of *APOL1* variants may be associated with other conditions beyond human African trypanosomiasis and CKD. We assessed the association of *APOL1* variants in a phenome wide study of a large population from the UK Biobank, a large-scale biomedical database and research resource containing genetic, lifestyle, and health information from half a million participants from across the UK^16^. Using indicators of CKD as a covariate (alongside other appropriate covariates), we performed a phenome-wide screen. We show that G1 and G2 are not equivalent ‘high-risk’ variants and that the G1/G2 compound heterozygous genotype stands out as being particularly associated with deleterious outcomes in a far wider range of conditions than previously reported. In addition, detailed examination of association with CKD indicators revealed that G1/G1, G2/G2 and G1/G2 genotypes all display associations with distinct CKD-related phenotypes.

## Methods

### Study design and participants

The UK Biobank is a prospective cohort study of 502,460 adults aged 40 to 69 years at enrolment between 2006 and 2010 from 22 assessment centres across the United Kingdom^16^. At the baseline study visit, participants underwent nurse-led interviews and completed detailed questionnaires about medical history, medication use, sociodemographic factors, and lifestyle. Participants underwent a range of physical assessments and provided blood and urine at the baseline visit. The UK Biobank study was approved by the North-West Multi-Centre Research Ethics Committee, and all participants provided written informed consent.

As *APOL1* G1 and G2 are predominantly found in people with recent African ancestry, we selected UK Biobank participants on the basis of genetic evidence of African ancestry from previously reported principal component data (PC1 > 100 and PC2 > 0)^16^. The total number of participants with African ancestry included in this study was 10,179.

### Genotyping

*APOL1* genotypes were obtained from the UK Biobank which used a custom Affymetrix array for the G1 (rs73885319) and N264K (rs73885316) alleles, and obtained G2 (rs71785313) genotypes by imputation against the UK10K haplotype panel merged with the 1000 Genomes Project phase 3 genotypes for G2 (rs71785313)^16^.

### Identification of hospital inpatient diagnoses

Records for hospital inpatients and deaths were identified via UK Biobank tables ‘hesin_diag’ and ‘death_cause’. The International Classification of Diseases is a system of diagnostic codes for classifying factors relating to healthcare such as diseases, symptoms, abnormal findings, social circumstances, and external causes of injury. International Classification of Diseases, Tenth Revision (ICD-10)^17^ codes were used to identify hospital inpatient diagnoses. The UK Biobank contains ICD-10 codes at four levels of increasing disease specificity (Chapters (Supplementary Table 1), Level 1, Level 2, and Level 3). In order to identify associations with specific conditions, phenome-wide analysis was performed on ICD-10 codes at Level 2 (identified by a letter followed by two digits) and Level 3 (a letter followed by three digits).

### Definitions of chronic kidney disease and associated conditions

Indicators of CKD at UK Biobank enrolment were defined in accordance with guidelines from the Kidney Disease: Improving Global Outcomes clinical practice guidelines^18^: either an elevated urinary albumin:creatinine ratio (uACR > 3 mg/mmol), or decreased estimated glomerular filtration rate (eGFR < 60 mL/min/1.73m^2^). eGFR at enrolment was calculated using the CKD-EPI 2021 creatinine and creatinine-cystatin C equations^19^ (which are not adjusted for race), using data collected by the UK Biobank at registration: age when attended assessment centre, sex, creatinine, and cystatin C. In all analyses, CKD identified by low eGFR means that eGFR was < 60 by either one or both of the two equations. Urinary albumin:creatinine ratio at enrolment was calculated using UK Biobank data fields for microalbumin in urine and creatinine (enzymatic) in urine. End stage kidney disease (ESKD) as of September 2022 was defined as reaching CKD stage G5 or the requirement for kidney replacement therapy, using ICD-10 codes or Office of Population Censuses and Surveys Classification of Surgical Operations and Procedures, Version 4 (OPCS4) codes as described in the UK Biobank’s Definitions of End Stage Renal Disease^20^ (Supplementary data: Identification of end stage kidney disease). Participants were considered hypertensive on UK Biobank registration if they met at least one of the following criteria: self-reported hypertension; prescription of one or more antihypertensive medications for cholesterol, blood pressure, diabetes, or taking exogenous hormones; recording during registration of a systolic blood pressure of > 140 mmHg; recording during registration of diastolic blood pressure of > 90 mmHg. Participants were considered to have diabetes on UK Biobank registration if they self-reported diabetes or were taking one or more of the following diabetes medications: insulin, gliclazide, glimepiride, tolbutamide, pioglitazone, rosiglitazone, repaglinide, or nateglinide.

### Covariates

For the phenome-wide screen, age, sex, body mass index, Townsend deprivation index, UK Biobank principal components 1 to10, and CKD (*i*.*e*. elevated albumin:creatinine ratio, **or** decreased glomerular filtration rate **or** algorithmically-defined end stage kidney disease) were selected as covariates. For examining associations with CKD and infectious diseases covariates were selected based on previously-identified risk factors for each condition (Supplementary Table 3). CKD was included as a covariate in order to identify associations with *APOL1* genotypes that were not mediated by kidney disease.

### Association analysis

The primary exposure variables were the six observed combined G0, G1 and G2 *APOL1* genotypes (Table 1). Only ICD-10 codes which were assigned to at least 30 participants were retained for analysis. Conditions which affected less than 30 participants were excluded from the analysis since there would be limited power to detect associations with rarer genotypes such as G2/G2. Firth’s bias-reduced logistic regression as implemented in R^21^ was used to control for separation since numbers of some genotypes are expected to be low in some tests, particularly where small numbers of participants had a particular condition. All statistical tests were 2-sided, where a p < 0.05 was considered statistically significant for the primary outcome.

**Table 1:**
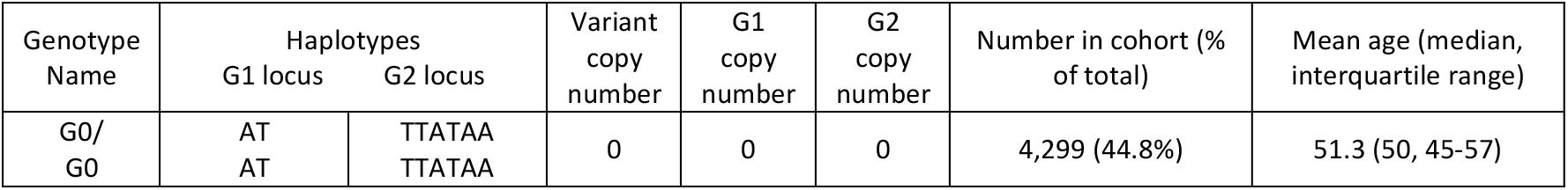
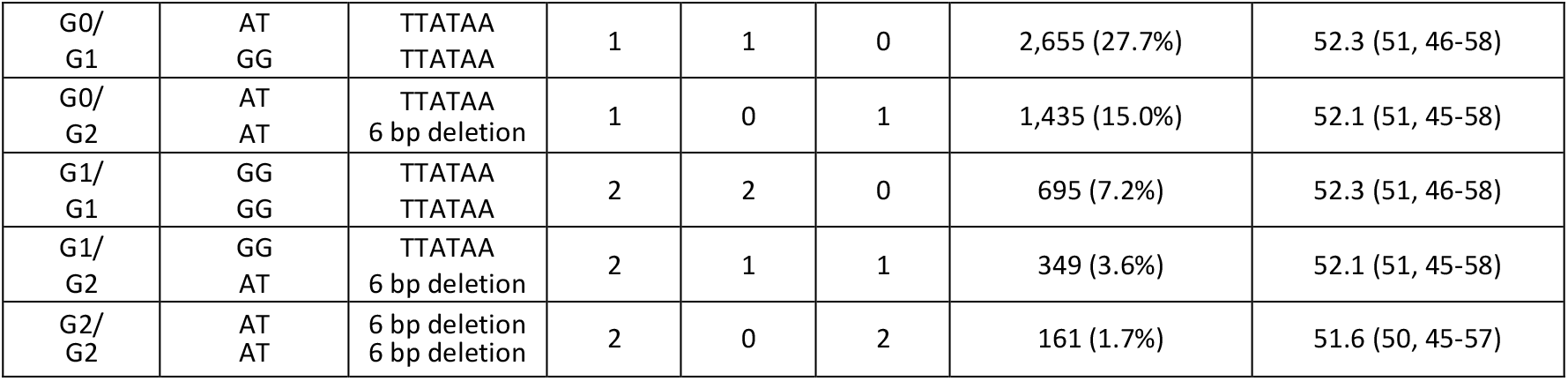
**Haplotype frequencies at the *APOL1* G1 and G2 loci in the UK Biobank cohort (n = 9,594). Genotypes with G1 and G2 on the same haplotype are theoretically possible but have not been observed.**

| Genotype Name | Haplotypes |  | Variant copy number | G1 copy number | G2 copy number | Number in cohort (% of total) | Mean age (median, interquartile range) |
| --- | --- | --- | --- | --- | --- | --- | --- |
|  | G1 locus | G2 locus |  |  |  |  |  |
| G0/G0 | AT<br>AT | TTATAA<br>TTATAA | 0 | 0 | 0 | 4,299 (44.8%) | 51.3 (50, 45-57) |
| G0/<br>G1 | AT<br>GG | TTATAA<br>TTATAA | 1 | 1 | 0 | 2,655 (27.7%) | 52.3 (51, 46-58) |
| G0/<br>G2 | AT<br>AT | TTATAA<br>6 bp deletion | 1 | 0 | 1 | 1,435 (15.0%) | 52.1 (51, 45-58) |
| G1/<br>G1 | GG<br>GG | TTATAA<br>TTATAA | 2 | 2 | 0 | 695 (7.2%) | 52.3 (51, 46-58) |
| G1/<br>G2 | GG<br>AT | TTATAA<br>6 bp deletion | 2 | 1 | 1 | 349 (3.6%) | 52.1 (51, 45-58) |
| G2/<br>G2 | AT<br>AT | 6 bp deletion<br>6 bp deletion | 2 | 0 | 2 | 161 (1.7%) | 51.6 (50, 45-57) |

The primary objective of this analysis was to identify which of the six observed *APOL1* compound genotypes had an excess of associations with ICD-10 codes, rather than demonstrating associations with any specific condition. For this purpose, a relatively relaxed false discovery rate (FDR) of 20% was chosen rather than a Bonferroni correction, because although particular conditions which pass the threshold may be false positives, the FDR estimates the excess of false positives compared to that expected by chance. The Qvalue^22^ package in R was used to calculate FDR.

## Results

### Identification of the cohort

Using previously-reported principal component data^16^, we identified 10,179 UK Biobank participants as having recent African ancestry. Of these, 9,594 had complete, unambiguous *APOL1* genotype data and were included in this study. This cohort accounted for 100% of participants with G1/G1, G1/G2, or G2/G2 genotypes, 95% of participants who were G0/G1, and 94% of those who were G0/G2. Genotype frequencies for the cohort are shown in Table 1. The allele frequencies in the cohort for G0, G1, and G2 were 66%, 23%, and 11%, respectively.

### Phenome-wide associations with *APOL1* variants

In order to identify which, if any, of the six observed *APOL1* genotypes had associations with diseases, we examined all hospital inpatient diagnoses as defined by ICD-10 codes. The UK Biobank contains ICD-10 codes at four levels (Chapter, Level 1, Level 2, and Level 3) of increasing disease specificity. We excluded those ICD-10 codes that affected fewer than 30 participants from the analysis since there would be very limited statistical power to detect associations between rarer *APOL1* genotypes and such conditions. We identified 470 Level 2 ICD-10 codes and 630 Level 3 ICD-10 codes that were recorded for at least 30 cohort members. Five models of association were examined: (1) whether conditions were associated with any of the five *APOL1* risk variant genotypes; (2) whether conditions were associated with G1 in either a dominant or a recessive model; (3) whether conditions were associated with G2 in either a dominant or a recessive model; (4) whether G1 and G2 were equivalent, and conditions were associated with carriage of either risk allele in either a dominant or a recessive model; (5) whether associations with either risk allele were dose-dependent, and therefore stronger with an increasing number of risk alleles (Table 2).

**Table 2:**
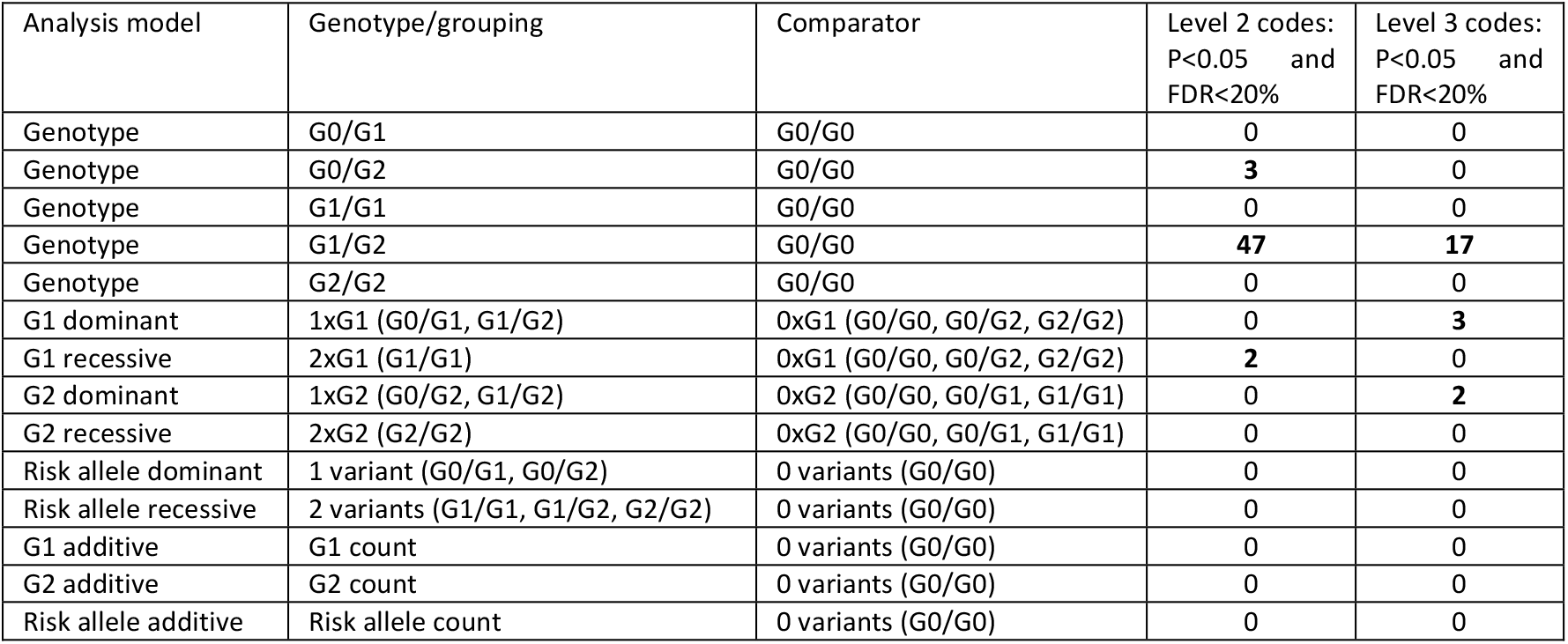
**Models of association considered in the phenome-wide screen, and number of potential associations identified by each model. Dominant models were where one risk allele was sufficient to produce phenotype, recessive alleles where two risk alleles were required to produce phenotype, and additive alleles (proportional to the number of risk alleles present).**

| Analysis model | Genotype/grouping | Comparator | Level 2 codes:<br>P<0.05 and<br>FDR<20% | Level 3 codes:<br>P<0.05 and<br>FDR<20% |
| --- | --- | --- | --- | --- |
| Genotype | G0/G1 | G0/G0 | 0 | 0 |
| Genotype | G0/G2 | G0/G0 | 3 | 0 |
| Genotype | G1/G1 | G0/G0 | 0 | 0 |
| Genotype | G1/G2 | G0/G0 | 47 | 17 |
| Genotype | G2/G2 | G0/G0 | 0 | 0 |
| G1 dominant | 1xG1 (G0/G1, G1/G2) | 0xG1 (G0/G0, G0/G2, G2/G2) | 0 | 3 |
| G1 recessive | 2xG1 (G1/G1) | 0xG1 (G0/G0, G0/G2, G2/G2) | 2 | 0 |
| G2 dominant | 1xG2 (G0/G2, G1/G2) | 0xG2 (G0/G0, G0/G1, G1/G1) | 0 | 2 |
| G2 recessive | 2xG2 (G2/G2) | 0xG2 (G0/G0, G0/G1, G1/G1) | 0 | 0 |
| Risk allele dominant | 1 variant (G0/G1, G0/G2) | 0 variants (G0/G0) | 0 | 0 |
| Risk allele recessive | 2 variants (G1/G1, G1/G2, G2/G2) | 0 variants (G0/G0) | 0 | 0 |
| G1 additive | G1 count | 0 variants (G0/G0) | 0 | 0 |
| G2 additive | G2 count | 0 variants (G0/G0) | 0 | 0 |
| Risk allele additive | Risk allele count | 0 variants (G0/G0) | 0 | 0 |

Each phenotype was tested for an association with *APOL1* genotype using logistic regression with age, sex, body mass index, Townsend deprivation index, evidence of chronic kidney disease (eGFR < 60 mL/min/1.73m^2^ or uACR > 3 mg/mmol or algorithmically-defined ESKD), and UK Biobank principal components 1-10 used as covariates (Supplementary Table 3). A false discovery rate was used to adjust for multiple testing. The aim of the study was to identify the haplotype combinations which had an excess of associations with ICD-10 codes, rather than demonstrating associations with any specific condition, making it appropriate to use a relatively relaxed FDR threshold of 20%, in a similar approach to other association studies^23,24^. Seventy-four potential associations (p < 0.05 and FDR < 20%) were detected (Supplementary Tables 4 and 5) across 52 Level 2 ICD-10 codes and 22 Level 3 ICD-10 codes (Table 2). One specific risk compound genotype dominated this analysis, with 64 (86.5%) of the associations linked to the G1/G2 genotype (Figure 1): particularly remarkable given that 695 and 349 participants had G1/G1 and G1/G2 genotypes respectively, indicating a substantially greater power to detect associations with G1/G1 than G1/G2, but that these were not found (Supplementary data: Sensitivity Analysis). Conversely, there were only 161 participants with the G2/G2 genotype (Table 1), and it is possible that the complete absence of associations with G2/G2 with FDR < 20% is due to lack of statistical power (Supplementary data: Sensitivity Analysis).

**Figure 1:**
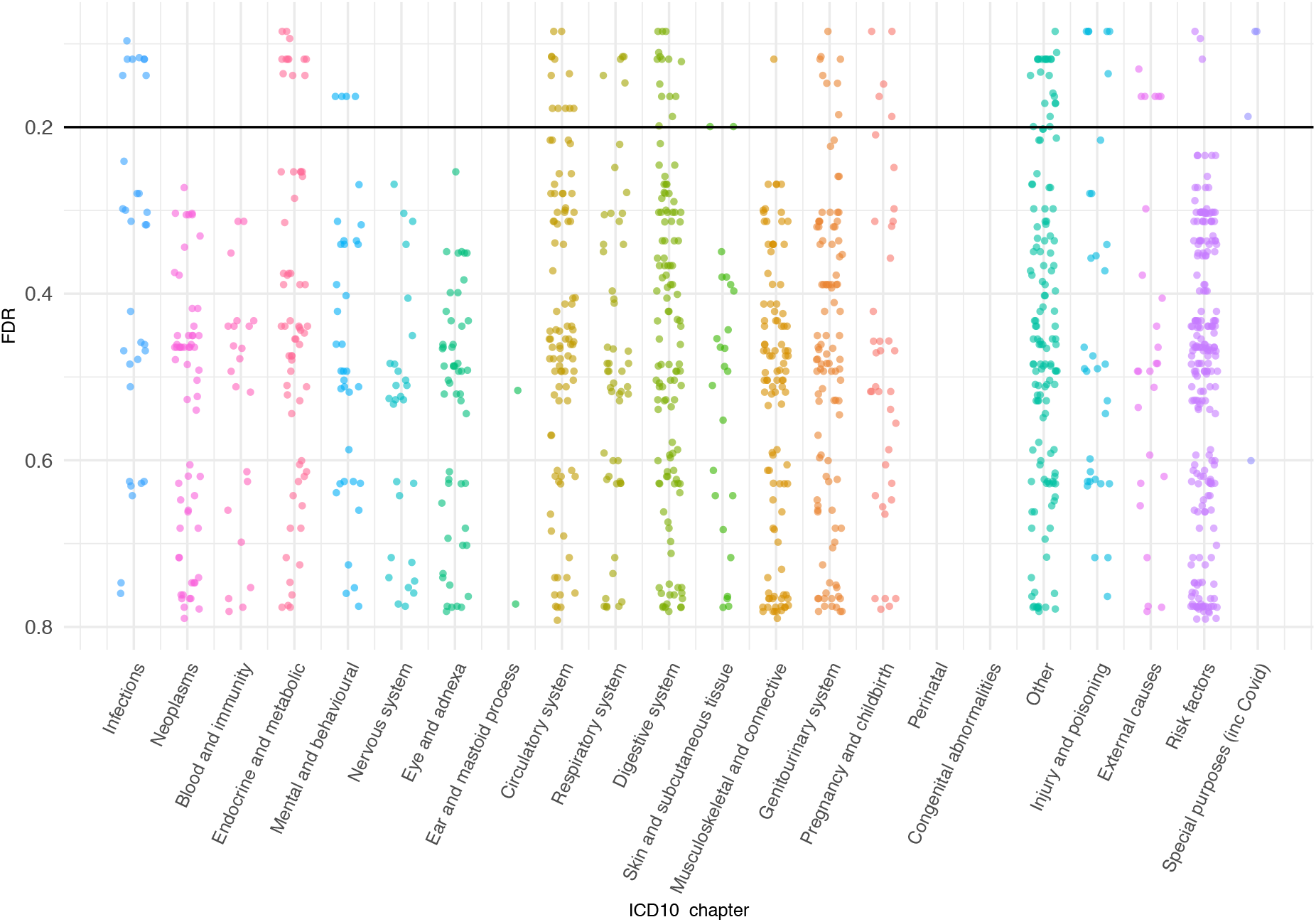
False discovery rate values for association between Level 2 and Level 3 ICD-10 codes and *APOL1* G1/G2 genotype. Horizontal line indicates the threshold that was used for the false discovery rate (20%) for a potentially significant association.

Non-G1/G2 associations are shown in Supplementary Tables 4 and 5 and Supplementary Figure 1. Notably, no potential associations were detected for the high-risk variants when considered collectively. Odds ratios indicated that 72 of the 74 potential associations (including all potential associations with the G1/G2 genotype) were deleterious, with increased rates of hospitalisation among cohort members with the variant genotype/grouping.

We then used z-score tests to examine whether the 64 associations with G1/G2 genotype were randomly distributed across ICD-10 codes, or whether specific ICD-10 coding chapters were over- or under-represented. ICD-10 chapters that were over-represented were; chapters I (certain infectious and parasitic diseases) with association detected with 6/29 codes (21%, p = 0.001), chapter IV (endocrine, nutritional, and metabolic diseases) with association detected with 7/50 codes (14.0%, p = 0.02), and chapter XXII (codes for special purposes (such as new diseases of uncertain aetiology), which includes the U071 code for the identification of COVID-19) association detected in 4/5 codes (80%, p = 6×10^−5^) (Supplementary Table 6). No coding chapters were significantly under-represented.

Finally, we tested whether any of the *APOL1* genotypes were associated with an excess of ICD-10 codes. Each UK Biobank participants in our cohort had a mean of 9.96 ICD-10 Level 3 codes assigned to them. In this analysis the G1/G2 genotype again emerged as distinct with a significantly higher average number of ICD-10 Level 3 codes (12.48) than that recorded for participants with the G0/G0 genotype (9.52, p=0.00006). No such differences were observed for any other *APOL1* genotypes containing risk variants (Supplementary Table 7).

### Associations between *APOL1* G1/G2 and hospitalisation due to infectious diseases

The phenome-wide screen indicated an association between the G1/G2 genotype and a range of hospital inpatient diagnoses, with an overrepresentation of codes related to infectious diseases (chapters I and XXII), including COVID-19 (ICD-10 code U071). Chi-squared tests confirmed that the G1/G2 genotype was not associated with SARS-CoV-2 testing (p = 0.08) or positivity (p = 0.32). To test if G1/G2 was associated with hospitalisation or death in G0/G0 and G1/G2 genotypes. Covariates used were previously-associated COVID-19 risk factors^25–27^ (age, sex, CKD, atrial fibrillation, depression, chronic obstructive pulmonary disorder, dementia, type 2 diabetes, body mass index, Townsend deprivation index, and UK Biobank principal components 1-10) (Supplementary Table 3). Associations were detected for G1/G2 with hospitalisation (OR = 2.4, 95% CI: 1.3-4.3, p = 0.008) and death (OR = 7.3, 95% CI: 2.7-19.5, p = 0.0002) (Supplementary Table 8). No association with poor outcomes in COVID-19 were detected for the other genotypes: G0/G1, G0/G2, G1/G1, or G2/G2.

We re-examined the associations we identified in the phenome-wide screen between G1/G2 and poor outcomes in other infectious diseases (*i*.*e*., viral pneumonia, gram-negative bacterial sepsis, diarrhoea/gastroenteritis and viral hepatitis; Supplementary Tables 4 and 5), adjusting the covariates as appropriate for each disease^28–31^ (Supplementary Table 3). Associations were detected for hospitalisation due to viral pneumonia (OR = 2.4, 95% CI: 1.2-4.6, p = 0.01), gram-negative bacterial sepsis (OR = 4.3, 95% CI: 1.4-11.9, p = 0.01), and diarrhoea/gastroenteritis of presumed infectious origin (OR = 1.9, 95% CI: 1.1-3.0, p = 0.02) (Supplementary Table 8).

### Associations between *APOL1* variants and chronic kidney disease

Data from the phenome-wide screen demonstrated that different *APOL1* genotypes had distinct patterns of association. We therefore tested whether these differences also applied to CKD: the disease in which associations between *APOL1* genotypes and pathology were first discovered. Within the cohort, 973 individuals (10.1%) were identified as having at least one indicator of CKD on UK Biobank registration (*i*.*e*., eGFR (creatinine) < 60 mL/min/1.73m^2^ **or** eGFR (creatinine-cystatin C) < 60 mL/min/1.73m^2^ **or** uACR > 3 mg/mmol) (Supplementary Table 9). In the following analyses, individuals with eGFR < 60 mL/min/1.73m^2^ by either the creatinine or the creatinine-cystatin C equation were considered to have CKD. Both eGFR and uACR indicators of CKD were present in 85 individuals (0.9% overall, 8.7% of those with CKD). We investigated the association between carriage of two high-risk variants (genotypes G1/G1, G1/G2, and G2/G2) in our UK Biobank cohort, in a similar manner to previous studies on African Americans^2^, using the first 10 UK Biobank principal components as covariates along with previously-associated CKD risk factors^32^ (age, sex, body mass index, Townsend deprivation index, hypertension, and diabetes). Associations were detected between carriage of two high-risk variants and CKD as defined as either uACR > 3 mg/mmol or eGFR < 60 mL/min/1.73m^2^, as well as the two factors individually (Table 3). Recent evidence suggests that an additional *APOL1* variant, N264K (rs73885316) reduces the cytotoxicity of *APOL1* high-risk variants in HEK293 kidney cells^33^. This variant was present in 294 cohort members (3.1%) however when it was applied as an additional covariate, it did not affect associations reported here. The N264K variant alone was not associated with any CKD phenotypes examined in this study (data not shown).

**Table 3:** **Association of risk indicators of chronic kidney disease with number of *APOL1* risk variants, compared to 0 variants, adjusted for age, sex, body mass index, diabetes, hypertension, Townsend deprivation index, and UK Biobank genetic principal components 1-10. Genotypes with p values < 0.05 are shown in bold. The numbers and percentages are shown in Supplementary Table 9.**

| Genotype | n (total) | uACR > 3 mg/mmol or<br>eGFR < 60<br>mL/min/1.73m<sup>2</sup> |  | uACR > 3 mg/mmol |  | eGFR < 60<br>mL/min/1.73m<sup>2</sup> |  |
| --- | --- | --- | --- | --- | --- | --- | --- |
|  |  | Odds ratio<br>(95% CI) | p | Odds ratio<br>(95% CI) | P | Odds ratio<br>(95% CI) | P |
| 0 variants | 4,299 | 1.0 (ref) |  | 1.0 (ref) |  | 1.0 (ref) |  |
| 1 variant | 4,090 | 1.1 (0.9-1.3) | 0.43 | 1.1 (0.9-1.3) | 0.37 | 1.0 (0.7-1.3) | 0.87 |
| 2 variants | 1,205 | **1.5 (1.2-1.8)** | **0.0006** | **1.5 (1.1-1.9)** | **0.004** | **1.5 (1.1-2.2)** | **0.02** |

We then examined each genotype combination individually, using the same statistical method. First, we demonstrated that the inclusion of *APOL1* G1 and G2 combined genotypes in a logistic regression model significantly improved the fit of the model to the data (ANOVA p=0.0096) and that therefore *APOL1* genotypes contribute to risk of CKD. We tested whether the individual combined genotypes made equivalent contributions to risk of CKD or whether it was important to examine them separately by constraining all alternate genotypes to have the same effect and comparing this model with the full model with one reference genotype and five alternate genotypes. The full model was a significantly better fit to the data (ANOVA p = 0.018), demonstrating that examining individual genotypes is more informative than grouping all two-variant genotypes together. Logistic regression using individual genotype combinations showed that G1/G1 and G1/G2 were associated with having an indicator of CKD at UK Biobank registration (either uACR > 3 mg/mmol or eGFR < 60 mL/min/1.73m^2^), however no such association was detected for G2/G2 (Table 4). Considering indicators of CKD individually, having uACR > 3 mg/mmol was associated specifically with G1/G1 while eGFR < 60 mL/min/1.73m^2^ was associated with G1/G2, and G2/G2. By examining each two-risk-variant genotype individually rather than grouping them together, we detect complexities in the relationship between *APOL1* risk variants and CKD, suggesting that APOL1-mediated cell injury in CKD might be a result of more than one genotype-specific molecular pathway.

**Table 4:** **Association of risk indicators of chronic kidney disease with *APOL1* genotypes compared to G0/G0, adjusted for age, sex, body mass index, diabetes, hypertension, Townsend deprivation index, and UK Biobank genetic principal components 1-10. Genotypes with p values < 0.05 are shown in bold. The numbers of affected participants with each genotype and percentages are shown in Supplementary Table 10.**

| Genotype | n (total) | uACR > 3 mg/mmol or<br>eGFR < 60 mL/min/1.73m<sup>2</sup> |  | uACR > 3 mg/mmol |  | eGFR < 60<br>mL/min/1.73m<sup>2</sup> |  |
| --- | --- | --- | --- | --- | --- | --- | --- |
|  |  | Odds ratio<br>(95% CI) | p | Odds ratio<br>(95% CI) | P | Odds ratio<br>(95% CI) | P |
| G0/G0 | 4,299 | 1.0 (ref) |  | 1.0 (ref) |  | 1.0 (ref) |  |
| G0/G1 | 2,655 | 1.0 (0.9-1.2) | 0.72 | 1.0 (0.8-1.3) | 0.67 | 0.9 (0.7-1.3) | 0.65 |
| G0/G2 | 1,435 | 1.1 (0.9-1.4) | 0.25 | 1.2 (0.9-1.5) | 0.20 | 1.1 (0.7-1.5) | 0.70 |
| **G1/G1** | **695** | **1.5 (1.1-1.9)** | **0.004** | **1.6 (1.2-2.2)** | **0.002** | 1.3 (0.8-2.0) | 0.29 |
| **G1/G2** | **349** | **1.6 (1.1-2.2)** | **0.01** | 1.3 (0.9-2.0) | 0.15 | **1.8 (1.0-2.9)** | **0.04** |
| **G2/G2** | 161 | 1.2 (0.7-2.0) | 0.51 | 1.1 (0.5-2.0) | 0.88 | **2.6 (1.3-4.9)** | **0.01** |

### Associations between *APOL1* variants and end stage kidney disease

Having identified genotypic associations with indicators of CKD measured at the time of participants’ registration to the UK Biobank, we examined incidences of end stage kidney disease (ESKD) in our cohort recorded by September 2022. ESKD was defined algorithmically according to the UK Biobank’s guidelines^20^, and based on ICD-10 and OPCS4 codes recorded for each cohort member. There was strong correlation with subsequent ESKD with having eGFR < 60 mL/min/1.73m^2^ (OR = 23.2, 95% CI: 13.6-40.0, p < 1×10^−12^) or uACR > 3 mg/mmol (OR = 6.6, 95% CI: 4.0-11.0, p = 2×10^−12^) at UK Biobank recruitment. We then examined genotypic associations with ESKD: an association was detected solely for the G1/G2 genotype (OR = 1.9, 95% CI: 1.9-2.8, p = 0.004) (Table 5).

**Table 5:** **Association of risk of end stage kidney disease with *APOL1* genotypes compared to G0/G0, adjusted for age, sex, body mass index, diabetes, hypertension, Townsend deprivation index, and UK Biobank genetic principal components 1-10. P values < 0.05 are shown in bold.**

| Genotype | n (total) | n (end stage kidney disease) (%) | Odds ratio | P |
| --- | --- | --- | --- | --- |
| G0/G0 | 4,299 | 31 (0.7%) |  |  |
| G0/G1 | 2,655 | 20 (0.8%) | 1.0 (0.6-1.9) | 0.94 |
| G0/G2 | 1,435 | 11 (0.8%) | 1.1 (0.5-2.1) | 0.88 |
| G1/G1 | 695 | 9 (1.3%) | 1.5 (0.7-3.2) | 0.30 |
| **G1/G2** | **349** | **10 (2.9%)** | **3.4 (1.5-7.2)** | **0.004** |
| G2/G2 | 161 | 1 (0.6%) | 1.3 (0.2-5.3) | 0.75 |

In analysing the relationship between *APOL1* genotypes and CKD, we identified associations that were not apparent from treating G1 and G2 as equivalent ‘high-risk’ alleles, and that the two-variant *APOL1* genotypes displayed distinct phenotypes in terms of CKD indicators and disease progression.

## Discussion

Relationships between G1 and G2 *APOL1* variants and several non-communicable diseases including chronic kidney disease are well-established in African American populations^1^. Here, for the first time, we confirm this association in a population-based cohort of people with recent African ancestry living in the United Kingdom. We show that one specific multi-locus genotype, G1/G2, is associated with multiple disease phenotypes with primarily deleterious outcomes, and that the scope of conditions affected by this particular genotype might be far wider than previously thought. While studies of the relationship between high-risk *APOL1* variants and CKD have largely focused on the number of variants in an individual, with carriage of two variants considered deleterious, we provide evidence that the different combinations of *APOL1* variants are associated with different phenotypes.

Our phenome-wide association study revealed 74 potential associations, 72 of which were deleterious: far more than would be expected if the potential associations occurred by chance (p = 1×10^−11^). No association was detected when grouping G1/G1, G1/G2 and G2/G2 risk genotypes together. However, when each genotype was examined individually, a spectrum of potential *APOL1*-associated conditions was detected. This revealed that G1/G2 is potentially associated with several disease phenotypes, and that its effect is often masked by the lack of association with G1/G1 and G2/G2. The lack of association when grouping risk genotypes together is consistent with a similar phenome-wide association study of high-risk *APOL1* variants that was conducted using data from 6,579 African American participants from Penn Medicine Biobank and Vanderbilt BioVU^11^. When CKD/ kidney failure status (defined as eGFR <60 mL/min/1.73m^2^ or at least two ICD codes for dialysis or kidney transplant) was included as a covariate and a stringent Bonferroni correction applied, no non-renal diseases were associated when *APOL1* risk variants were grouped together.

Our analysis identified multiple associations with the G1/G2 genotype, despite the greater power to detect associations with the G1/G1 combination than the G1/G2 combination. Sixty-four associations with G1/G2 had FDR <20% were detected whereas no associations with G1/G1 had FDR <20% (Table 2): a statistically significant difference between the two genotypes (p < 2.2 × 10^−16^). Although there was less power to detect associations with the G2/G2 haplotype combination the zero associations with G2/G2 with FDR < 20% (Table 2) suggests that this combination may have less impact on disease that G1/G2. Further targeted studies will be required to identify which of these associations are true positives.

Infectious diseases are overrepresented in terms of conditions that we detected as being potentially associated with G1/G2. The risk of hospitalisation or death from COVID-19 is disproportionately higher in people of African ancestry. There is growing evidence that carriage of two high-risk variant *APOL1* alleles is associated with adverse outcomes in COVID-19. In African Americans infected with SARS-CoV-2, carriage of two *APOL1* variants is associated with collapsing glomerulopathy^7^, acute kidney injury, persistent AKI, and requirement for kidney replacement therapy^8^. Among African Americans hospitalised with COVID-19, carriage of two high-risk *APOL1* variants has been associated with increased AKI severity and death^9^. When considering individual genotypes in our study we revealed that associations with these phenotypes were driven primarily by the G1/G2 genotype, and the inclusion of CKD as a covariate suggests a potential non-renal pathway through which G1/G2 affects SARS-CoV-2 infection outcome.

Infectious disease is a major evolutionary driving force of natural selection. The relationship between high-risk *APOL1* variants and human African trypanosomiasis is well-characterised: G2 is associated with protection from *T*.*b. rhodesiense* infection, but conversely is associated with increased disease severity of *T*.*b. gambiense*, whereas G1 is associated with milder disease severity in *T*.*b. gambiense*^3^. However, the modern-day distribution of G1 and G2 with both variants being at higher frequency in West Africa does not correspond to the current geographic ranges of the two parasite species: *T*.*b. rhodesiense* being found only in East Africa and *T*.*b. gambiense* in West and Central Africa. This suggests that factors other than *T. brucei* sub-species might have provided the selection pressure for the allele distribution. Infectious diseases such as malaria, cholera, dengue, and typhoid have affected millions of people in sub-Saharan Africa for centuries and may have driven the selection of *APOL1* variants but are either rare or absent from the cohort described here. As a result, it has not been possible to assess the impact that *APOL1* genotype has on such conditions or identify other conditions which might be involved in positive selection for *APOL1* variants, however this warrants further investigation. Carriage of two high-risk *APOL1* variants has been strongly associated with HIV-associated nephropathy (HIVAN) with odds ratio of 89^5^, but also associated with protection from HIV-associated opportunistic infections^34^. However, in our study we were unable to examine this association in our dataset as the UK biobank did not contain enough cases of opportunistic infections in HIV-infected individuals to make an assessment.

Previously, carriage of two high-risk variant alleles has been associated with sepsis^35^. In our study, we have identified a similar association driven primarily by G1/G2. We also extended the range of infections in which outcomes are associated with *APOL1* genotype (specifically G1/G2) to viral pneumonia, and gastroenteritis.

The phenome-wide screen also identified potential associations between G1/G2 and several conditions related to the transport of ions and other metabolites across membranes (including magnesium metabolisms disorders (OR = 7.1, 95% CI: 2.5-18.9, p = 0.0003), hyperkalaemia (OR = 3.4, 95% CI: 1.5-7.2, p = 0.001), hypoglycaemia (OR = 3.0, 95% CI:1.5-5.6, p = 0.002), and vitamin D deficiency (OR = 2.7, 95% CI: 1.5-4.5, p = 0.001): all of which are related to kidney function, however these associations continue to be detected when CKD is included as a covariate. APOL1 forms an ion channel: in human African trypanosomiasis, APOL1 forms pores in parasite membranes, disrupting ionic balance, and causing lysis^36^. Therefore, the associations with the transport of ions and metabolites are intriguing and is suggestive of a wider role for APOL1 channels.

With regards to CKD, we demonstrated that different genotypes were associated with different disease indicators. G1/G1 was associated with elevated uACR (>3 mg/mmol), G2/G2 was associated with decreased eGFR (< 60 mL/min/1.73m^2^), and G1/G2 is associated with decreased eGFR and more rapid progression to ESKD. Calculations of eGFR were performed using the CKD-EPI 2021 equations^19^, which are not adjusted for ancestry: improving accuracy and precision in estimating GFR for black adults^37^.

Recently, a similar association between G1/G1 and elevated uACR was also shown in a sub-Saharan African cohort^15^, however the authors did not detect an association with eGFR for any *APOL1* genotype. Several factors might account for this difference between studies, such as the use of GFR estimating equations having limited validation in sub-Saharan Africa^15^, or low GFR being influenced by other genetic or environmental factors that differentiate the UK population of African ancestry from sub-Saharan Africans. Notably, the majority (91.2%) of the cohort considered to have CKD in this study had just one indicator: *either* reduced eGFR *or* elevated uACR. The distinct associations observed for each two-high-risk-variant genotype might indicate that cell injury in CKD is modulated by multiple (potentially opposing) genotype-specific molecular pathways that each generate distinct metabolic signatures. An observation that might account for the multiple mechanisms that have been proposed for driving APOL1-related kidney damage^38^.

The membrane-damaging properties of APOL1 might be the common mechanism that explains both the destruction of parasites and kidney injury^38^, possibly affecting the multiple pathways that have previously been proposed including autophagy, lysosomal permeability, pyroptosis, mitochondrial dysfunction, impairment of vacuolar acidification, activation of stress-related kinases, endoplasmic reticulum stress, and mitophagy^35,39^. The number of subunits that comprise an APOL1 pore is unclear, however it has been proposed that the protein acts as a dimer^40^. One possible hypothesis to explain the wide diversity of diseases associated with *APOL1* G1/G2 is that pores made of heterogenous subunits are either not able to form pores, or form dysfunctional pores, disrupting cellular function and causing adverse phenotypes.

The increased disease burden in G1/G2 compound heterozygotes represents an epistatic interaction, which might explain why it has not been detected in previous genome-wide association studies (GWAS) that focus on additive associations with individual variants. Since the earliest GWAS, it has been clear that the loci that have been identified only account for a modest proportion of heritability. It has been proposed that this ‘missing heritability’ is due to difficulties in designing well-powered experiments: (1) sample size must be very large to detect loci with small effects; (2) combinations of variants might have joint effects larger than the sum of their individual effects – epistasis which would require an exponential increase in sample size to detect in a genome-wide search; (3) structural variants such copy number variation are not reliably detected by current genotyping methods; (4) problems with phenotyping^41^. This study provides an example of the potential for epistasis to explain part of the missing heritability. The discovery of an allelic interaction that is associated with a spectrum of human conditions has potentially far-reaching consequences: there are potentially many other such complex genotypes that impact on human health, and computational methods to simultaneously screen for associations with multiple combinations of alleles would be required.

Associations previously reported between *APOL1* risk alleles and conditions such as CKD have led to calls for the introduction of testing to identify *APOL1* genotype and to minimise the risk of kidney transplant failure. While the potential associations described here require confirmation in other cohorts, they indicate a wide spectrum of conditions that are associated with *APOL1* risk alleles, and particularly with the G1/G2 genotype. Previous studies have highlighted ethical considerations of such testing^42^, however strong support for testing has recently been reported among African Americans attending hypertension and nephrology clinics in the United States^43^, and disclosure of high-risk *APOL1* genotypes to hypertension patients has led to reductions in blood pressure and lifestyle changes such as improved dietary and exercise habits^44^. An affordable, rapid, point-of-care test for *APOL1* genotype would enable testing to be performed in both affluent and resource-poor settings and could have important implications at individual and population level by identifying those who would benefit from targeted early intervention and treatment.

The work described here is especially relevant to geographical regions where *APOL1* risk alleles are common such as West and Central Africa and the recent African diaspora which accounts for 140 million individuals worldwide^45^.

## Data Availability

This research has been conducted using the UK Biobank Resource under application number 66821. All bona fide researchers can apply to use the UK Biobank resource for health related research that is in the public interest.

https://www.ukbiobank.ac.uk

## Acknowledgements

This research has been conducted using data from the UK Biobank, a major biomedical database: www.ukbiobank.ac.uk.

## Approval

Access to the UK Biobank data was granted for this work under UK Biobank application number 66821.

## Funding

This study was funded by the Wellcome Trust (209511/Z/17/Z) and H3Africa (H3A/18/004).

## Declaration of interests

The authors have no conflicting interests.

## Notes

### Competing Interest Statement

The authors have declared no competing interest.

### Author Declarations

Access to the UK Biobank data was granted for this work under UK Biobank application number 66821.

