## Supplementary data for "Phenome-wide analysis of *APOL1* risk variants reveals associations between one combination of haplotypes and multiple disease phenotypes in addition to chronic kidney disease"

| **Chapter** | **ICD-10 codes** | **Description** |
| --- | --- | --- |
| I | A00-B99 | Certain infectious and parasitic diseases |
| II | C00-D48 | Neoplasms |
| III | D50-D89 | Diseases of the blood and blood-forming organs and certain disorders involving the immune mechanism |
| IV | E00-E90 | Endocrine, nutritional, and metabolic disorders |
| V | F00-F99 | Mental and behavioural disorders |
| VI | G00-G99 | Diseases of the nervous system |
| VII | H00-H59 | Diseases of the eye and adnexa |
| VIII | H60-H95 | Diseases of the ear and mastoid process |
| IX | I00-I99 | Diseases of the circulatory system |
| X | J00-J99 | Diseases of the respiratory system |
| XI | K00-K93 | Diseases of the digestive system |
| XII | L00-L99 | Diseases of the skin and subcutaneous tissue |
| XIII | M00-M99 | Diseases of the musculoskeletal system and connective tissue |
| XIV | N00-N99 | Diseases of the genitourinary system |
| XV | O00-O99 | Pregnancy, childbirth, and the puerperium |
| XVI | P00-P96 | Certain conditions of the perinatal period |
| XVII | Q00-Q99 | Congenital malformations, deformations, and clinical abnormalities |
| XVIII | R00-R99 | Symptoms, signs, and abnormal clinical and laboratory findings, not elsewhere classified |
| XIX | S00-T98 | Injury, poisoning, and certain other consequences of external causes |
| XX | V01-Y98 | External causes of morbidity and mortality |
| XXI | Z00-Z99 | Factors influencing health status and contact with health services |
| XXII | U00-U85 | Codes for special purposes |

**Supplementary Table 1: International Classification of Disease, Version 10 codes, by chapter. Adapted from World Health Organization *ICD-10 Version:2019*^17^.**

| **Data collected at UK Biobank enrolment** | **UK Biobank data field(s)** |
| --- | --- |
| Age when attended assessment centre | 21003 |
| Blood pressure, diastolic | 4079 |
| Blood pressure, systolic | 4080 |
| Creatinine | 30700 |
| Creatinine (enzymatic) in urine | 30510 |
| Cystatin C | 30720 |
| Medications taken | 6153, 6177, 20003 |
| Microalbumin in urine | 30500 |
| Self-reported illness | 20002 |
| Sex | 31 |
| Standing height | 50 |
| Townsend Deprivation Index | 189 |
| Weight | 21002 |

**Supplementary Table 2: UK Biobank data fields used to identify chronic kidney disease indicators, associated conditions, and covariates as described in Methods.**

| **Condition/analysis** | **Covariates** | **Reference(s)** |
| --- | --- | --- |
| Chronic kidney disease | Age, sex, body mass index, Townsend deprivation index, UK Biobank principal components 1-10, diabetes, hypertension | Luyckx *et al*., 2020 |
| Phenome-wide screening | Age, sex, body mass index, Townsend deprivation index, UK Biobank principal components 1-10, chronic kidney disease | N/A |
| Hospitalisation/death due to COVID-19 | Age, sex, body mass index, Townsend deprivation index, UK Biobank principal components 1-10, chronic kidney disease, atrial fibrillation, depression, chronic obstructive pulmonary disease, dementia, type 2 diabetes, | Docherty *et al*., 2020; Atkins *et al*., 2020; Woodward *et al*., 2021 |
| Hospitalisation due to viral pneumonia | Age, sex, body mass index, Townsend deprivation index, UK Biobank principal components 1-10, chronic kidney disease, chronic obstructive pulmonary disease, human immunodeficiency virus, cigarette smoking status, level of alcohol consumption | Torres *et al*., 2013 |
| Hospitalisation due to sepsis caused by gram-negative bacteria | Age, sex, body mass index, Townsend deprivation index, UK Biobank principal components 1-10, chronic kidney disease, cardiovascular disease, chronic level disease, chronic obstructive pulmonary disease | Drozd *et al*., 2021 |
| Hospitalisation due to chronic viral hepatitis | Age, sex, body mass index, Townsend deprivation index, UK Biobank principal components 1-10, chronic kidney disease, age of first sexual intercourse, number of sexual partners | Ecollan *et al*., 2020 |
| Hospitalisation due to gastroenteritis caused by an infectious agent | Age, sex, body mass index, Townsend deprivation index, UK Biobank principal components 1-10, chronic kidney disease, chronic liver disease, cardiovascular disease | Lavanchy *et al*., 2008 |

**Supplementary Table 3: Covariates used for regression analyses.**

| **Genotype/grouping** | **ICD-10 code** | **ICD-10 code descriptor** | **ICD-10 coding chapter** | **p-value** | **Odds ratio (95% CI)** | **False discovery rate** |
| --- | --- | --- | --- | --- | --- | --- |
| G0/G2 | H26 | Other cataract | VII | 0.001 | 1.4 (1.1-1.8) | 0.197 |
| G0/G2 | H35 | Other retinal disorders | VII | 0.0004 | 2.1 (1.4-3.1) | 0.175 |
| G0/G2 | I67 | Other cerebrovascular diseases | IX | 0.001 | 1.9 (1.3-2.8) | 0.197 |
| G1/G2 | A09 | Other gastroenteritis and colitis of infectious and unspecified origin | I | 0.01 | 2.0 (1.2-3.2) | 0.151 |
| G1/G2 | B18 | Chronic viral hepatitis | I | 0.006 | 4.1 (1.6-9.9) | 0.151 |
| G1/G2 | B96 | Other specified bacterial agents as the cause of diseases classified to other chapters | I | 0.02 | 1.8 (1.1-2.8) | 0.170 |
| G1/G2 | B97 | Viral agents as the cause of diseases classified to other chapters | I | 0.02 | 2.3 (1.1-4.3) | 0.170 |
| G1/G2 | E16 | Other disorders of pancreatic internal secretion | IV | 0.004 | 2.8 (1.4-5.3) | 0.125 |
| G1/G2 | E55 | Vitamin D deficiency | IV | 0.001 | 2.7 (1.5-4.5) | 0.095 |
| G1/G2 | E87 | Other disorders of fluid, electrolyte and acid-base balance | IV | 0.01 | 1.8 (1.1-2.7) | 0.167 |
| G1/G2 | G51 | Facial nerve disorders | VI | 0.009 | 5.9 (1.6-19.4) | 0.151 |
| G1/G2 | H81 | Disorders of vestibular function | VIII | 0.02 | 3.6 (1.2-9.3) | 0.184 |
| G1/G2 | I08 | Multiple valve diseases | IX | 0.02 | 2.5 (1.2-4.9) | 0.167 |
| G1/G2 | I11 | Hypertensive heart disease | IX | 0.02 | 5.3 (1.4-19.2) | 0.170 |
| G1/G2 | I35 | Nonrheumatic aortic valve disorders | IX | 0.008 | 3.7 (1.5-8.7) | 0.151 |
| G1/G2 | I45 | Right fascicular block | IX | 0.009 | 3.3 (1.4-7.2) | 0.151 |
| G1/G2 | I73 | Other peripheral vascular diseases | IX | 0.004 | 3.0 (1.5-5.9) | 0.125 |
| G1/G2 | I77 | Other disorders of arteries and arterioles | IX | 0.01 | 4.0 (1.4-10.0) | 0.152 |
| G1/G2 | J02 | Acute pharyngitis | X | 0.03 | 3.6 (1.2-9.5) | 0.192 |
| G1/G2 | J12 | Viral pneumonia, not elsewhere classified | X | 0.01 | 2.5 (1.3-4.8) | 0.152 |
| G1/G2 | J96 | Postprocedural respiratory disorders, not elsewhere classified | X | 0.009 | 2.3 (1.2-4.2) | 0.151 |
| G1/G2 | K22 | Other diseases of oesophagus | XI | 0.004 | 2.6 (1.4-4.7) | 0.125 |
| G1/G2 | K52 | Other noninfective gastroenteritis and colitis | XI | 0.03 | 1.8 (1.1-2.9) | 0.193 |
| G1/G2 | K56 | Paralytic ileus and intestinal obstruction without hernia | XI | 0.01 | 2.6 (1.2-4.9) | 0.155 |
| G1/G2 | K58 | Irritable bowel syndrome | XI | 0.02 | 2.3 (1.2-4.4) | 0.170 |
| G1/G2 | K59 | Other functional intestinal disorders | XI | 0.007 | 1.7 (1.2-2.5) | 0.151 |
| G1/G2 | K65 | Peritonitis | XI | 0.001 | 6.6 (2.2-19.4) | 0.095 |
| G1/G2 | K66 | Other disorders of peritoneum | XI | 0.009 | 2.8 (1.3-5.4) | 0.151 |
| G1/G2 | M15 | Polyarthrosis | XIII | 0.01 | 2.8 (1.3-5.6) | 0.167 |
| G1/G2 | N13 | Obstructive and reflux uropathy | XIV | 0.001 | 4.2 (1.8-8.9) | 0.095 |
| G1/G2 | N17 | Acute renal failure | XIV | 0.009 | 1.8 (1.2-2.7) | 0.151 |
| G1/G2 | N28 | Other disorders of kidney and ureter, not elsewhere classified | XIV | 0.01 | 2.4 (1.2-4.4) | 0.155 |
| G1/G2 | N32 | Other disorders of bladder | XIV | 0.02 | 1.9 (1.1-3.1) | 0.193 |
| G1/G2 | O26 | Maternal care for other conditions predominantly related to pregnancy | XV | 0.002 | 4.0 (1.7-8.5) | 0.095 |
| G1/G2 | O36 | Maternal care for other known or suspected fetal problems | XV | 0.03 | 2.8 (1.1-6.4) | 0.193 |
| G1/G2 | R00 | Abnormalities of heartbeat | XVIII | 0.02 | 1.7 (1.1-2.6) | 0.170 |
| G1/G2 | R06 | Abnormalities of breathing | XVIII | 0.02 | 1.7 (1.1-2.6) | 0.170 |
| G1/G2 | R10 | Abdominal and pelvic pain | XVIII | 0.02 | 1.4 (1.1-1.9) | 0.170 |
| G1/G2 | R42 | Dizziness and giddiness | XVIII | 0.003 | 2.3 (1.3-3.7) | 0.125 |
| G1/G2 | R51 | Headache | XVIII | 0.007 | 1.9 (1.2-2.9) | 0.151 |
| G1/G2 | R69 | Unknown and unspecified causes of morbidity | XVIII | 0.02 | 1.6 (1.1-2.3) | 0.184 |
| G1/G2 | T81 | Complications of procedures, not elsewhere classified | XIX | 0.005 | 1.9 (1.2-2.9) | 0.138 |
| G1/G2 | T83 | Complications of genitourinary prosthetic devices, implants and grafts | XIX | 0.02 | 2.9 (1.2-6.2) | 0.170 |
| G1/G2 | U07 | Emergency use of U07 | XXII | 0.002 | 2.6 (1.5-4.4) | 0.095 |
| G1/G2 | U82 | Resistance to betalactam antibiotics | XXII | 0.01 | 3.7 (1.3-9.1) | 0.167 |
| G1/G2 | U83 | Resistance to other antibiotics | XXII | 0.03 | 3.2 (1.2-7.9) | 0.193 |
| G1/G2 | Y84 | Other medical procedures as the cause of abnormal reaction of the patient, or of later complication, without mention of misadventure at the time of the procedure | XX | 0.02 | 2.6 (1.2-5.4) | 0.184 |
| G1/G2 | Z33 | Pregnant state, incidental | XXI | 0.02 | 5.1 (1.3-16.0) | 0.167 |
| G1/G2 | Z74 | Problems related to care-provider dependency | XXI | 0.03 | 3.8 (1.2-10.7) | 0.192 |
| G1/G2 | Z75 | Problems related to medical facilities and other health care | XXI | 0.001 | 4.5 (1.8-10.6) | 0.095 |
| 2 copies of G1 | J98 | Other respiratory disorders | X | 0.0009 | 0.3 (0.1-0.6) | 0.153 |
| 2 copies of G1 | Z95 | Presence of cardiac and vascular implants and grafts | XXI | 0.0005 | 0.3 (0.1-0.6) | 0.153 |

**Supplementary Table 4: Level 2 International Classification of Disease, Version 10 (ICD-10) codes for which a potential association with *APOL1* risk alleles was indicated by the phenome-wide screen using data from UK Biobank participants with African ancestry.**

| **Genotype/grouping** | **ICD-10 code** | **ICD-10 code descriptor** | **ICD-10 coding chapter** | **p-value** | **Odds ratio (95% CI)** | **False discovery rate** |
| --- | --- | --- | --- | --- | --- | --- |
| G1/G2 | A099 | Gastroenteritis and colitis of unspecified origin | I | 0.001 | 2.0 (1.2-3.2) | 0.151 |
| G1/G2 | A415 | Sepsis due to other Gram-negative organisms | I | 0.002 | 5.8 (2.0-15.4) | 0.138 |
| G1/G2 | E162 | Hypoglycaemia, unspecified | IV | 0.002 | 3.0 (1.5-5.6) | 0.138 |
| G1/G2 | E559 | Vitamin D deficiency, unspecified | **IV** | 0.001 | 2.7 (1.5-4.5) | 0.138 |
| G1/G2 | E834 | Disorders of magnesium metabolism | **IV** | 0.0003 | 7.1 (2.5-18.9) | 0.094 |
| G1/G2 | E875 | Hyperkalaemia | **IV** | 0.004 | 3.4 (1.5-7.2) | 0.138 |
| G1/G2 | I500 | Congestive heart failure | IX | 0.004 | 3.1 (1.5-6.0) | 0.138 |
| G1/G2 | I739 | Peripheral vascular disease, unspecified | IX | 0.002 | 4.1 (1.8-8.9) | 0.136 |
| G1/G2 | J969 | Respiratory failure, unspecified | X | 0.002 | 3.0 (1.5-5.6) | 0.138 |
| G1/G2 | K566 | Other and unspecified intestinal obstruction | XI | 0.0007 | 4.6 (2.0-10.0) | 0.121 |
| G1/G2 | K660 | Peritoneal adhesions | XI | 0.007 | 2.9 (1.4-5.8) | 0.187 |
| G1/G2 | N133 | Other and unspecified hydronephrosis | **XIV** | 0.004 | 5.7 (1.8-16.1) | 0.138 |
| G1/G2 | O034 | Spontaneous abortion : incomplete, without complication | **XV** | 0.007 | 5.1 (1.6-14.0) | 0.187 |
| G1/G2 | R073 | Other chest pain | **XVIII** | 0.004 | 2.1 (1.3-3.2) | 0.138 |
| G1/G2 | T810 | Haemorrhage and haematoma complicating a procedure, not elsewhere classified | **XIX** | 0.002 | 2.8 (1.5-5.0) | 0.136 |
| G1/G2 | U071 | COVID-19, virus identified | **XXII** | 0.007 | 2.4 (1.3-4.3) | 0.187 |
| G1/G2 | Z751 | Person awaiting admission to adequate facility elsewhere | **XXI** | 0.0004 | 7.5 (2.6-21.1) | 0.094 |
| 1 copy of G1 | E834 | Disorders of magnesium metabolism | XI | 0.0007 | 2.9 (1.6-5.6) | 0.199 |
| 1 copy of G1 | F101 | Mental and behavioural disorders due to use of alcohol : harmful use | V | 0.0002 | 2.4 (1.5-3.9) | 0.097 |
| 1 copy of G1 | Y836 | Removal of other organ (partial) (total) | XX | 0.001 | 1.7 (1.2-2.4) | 0.097 |
| 2 copies of G2 | O034 | Spontaneous abortion : incomplete, without complication | **XV** | 0.0004 | 3.1 (1.7-5.5) | 0.158 |
| 2 copies of G2 | Z751 | Person awaiting admission to adequate facility elsewhere | **XXI** | 0.0005 | 3.6 (1.8-7.4) | 0.158 |

**Supplementary Table 5: Level 3 International Classification of Disease, Version 10 (ICD-10) codes for which a potential association with *APOL1* risk alleles was indicated by the phenome-wide screen using data from UK Biobank participants with African ancestry.**

| **Chapter** | **Level 2 and 3 ICD-10 codes analysed** | **Level 2 and 3 ICD-10 codes with P<0.05 and FDR<20% (%)** | **p** |
| --- | --- | --- | --- |
| **I** | **29** | **6 (20.7%)** | **0.001** |
| II | 51 | 0 (0%) | 0.08 |
| III | 20 | 0 (0%) | 0.27 |
| **IV** | **50** | **7 (14.0%)** | **0.02** |
| V | 31 | 0 (0%) | 0.17 |
| VI | 31 | 1 (3.2%) | 0.54 |
| VII | 42 | 0 (0%) | 0.11 |
| VIII | 4 | 1 (25.0%) | 0.10 |
| IX | 87 | 8 (9.2%) | 0.20 |
| X | 50 | 4 (8.0%) | 0.52 |
| XI | 109 | 9 (8.3%) | 0.31 |
| XII | 23 | 0 (0%) | 0.23 |
| XIII | 87 | 1 (1.1%) | 0.07 |
| XIV | 96 | 5 (5.2%) | 0.80 |
| XV | 40 | 3 (7.5%) | 0.66 |
| XVI | 0 | 0 (0%) | N/A |
| XVII | 0 | 0 (0%) | N/A |
| XVIII | 122 | 7 (5.7%) | 0.97 |
| XIX | 34 | 3 (8.8%) | 0.47 |
| XX | 29 | 1 (3.4%) | 0.59 |
| XXI | 160 | 4 (2.5%) | 0.08 |
| **XXII** | **5** | **4 (80%)** | **0.00006** |
| Total | 1100 | 64 (5.8%) |  |

**Supplementary Table 6: Counts of Level 2 and Level 3 ICD-10 codes for which a potential association with the G1/G2 genotype was indicated by the phenome-wide screen, by ICD-10 chapter. P values are for an excess of phenotypes with an association in each chapter calculated using z-score tests.**

| **Genotype** | **n (total)** | **ICD-10 codes per participant** | **p** |
| --- | --- | --- | --- |
| G0/G0 | 4,299 | 9.52 |  |
| G0/G1 | 2,655 | 10.47 | 0.14 |
| G0/G2 | 1,435 | 9.85 | 0.64 |
| G1/G1 | 695 | 9.90 | 0.89 |
| **G1/G2** | **349** | **12.48** | **0.0002** |
| G2/G2 | 161 | 9.27 | 0.43 |

**Supplementary Table 7: International Classification of Disease, Version 10 (ICD-10) codes per participant, comparing *APOL1* genotypes containing risk variants relative to G0/G0. Adjusted for age, sex, body mass index, Townsend deprivation index, and UK Biobank genetic principal components 1-10. P values ≤0.05 are shown in bold.**

| **Phenotype** | **Relevant ICD-10 codes** | **n (G0/G0)** | **n (G1/G2)** | **Odds ratio**  **(95% CI)** | **p** |
| --- | --- | --- | --- | --- | --- |
| Hospitalisation due to COVID-19 | U071 | 60 (1.4%) | 15 (4.3%) | 2.4 (1.3-4.3) | 0.008 |
| Death due to COVID-19 | J100, J110, J12 | 15 (0.3%) | 8 (2.3%) | 7.3 (2.7-19.5) | 0.0002 |
| Hospitalisation due to viral pneumonia | J100, J110, J12 | 49 (1.1%) | 12 (3.4%) | 2.4 (1.2-4.6) | 0.01 |
| Hospitalisation due to gram-negative bacterial sepsis | A413, A415 | 16 (0.4%) | 6 (1.7%) | 4.3 (1.4-11.9) | 0.01 |
| Hospitalisation due to infectious origin gastroenteritis | A09 | 146 (3.4%) | 21 (6.0%) | 1.9 (1.1-3.0) | 0.02 |
| Hospitalisation due to chronic viral hepatitis | B18 | 26 (0.6%) | 8 (2.3%) | 3.1 (0.6-12.4) | 0.17 |

**Supplementary Table 8: Risk of hospitalisation or death due to infectious diseases indicated by the phenome-wide screen as being potentially associated, for UK Biobank participants with African ancestry, comparing genotype G1/G2 (n = 349) to G0/G0 (n = 4,299). For columns n (G0/G0) and n (G1/G2), figures in brackets indicate the percentage of the cohort displaying the phenotype being examined. Covariates used are as described in Supplementary Table 3.**

| **Number of variants** | **n (total)** | **uACR >3 mg/mmol or eGFR <60 mL/min/1.73m^2^** | **uACR >3 mg/mmol** | **eGFR <60 mL/min/1.73m^2^** |
| --- | --- | --- | --- | --- |
| 0 variants | 4,299 | 388 (9.0%) | 302 (7.0%) | 115 (2.7%) |
| 1 variant | 4,090 | 416 (10.2%) | 324 (7.9%) | 126 (3.1%) |
| 2 variants | 1,205 | 169 (14.0%) | 131 (10.9%) | 60 (5.0%) |

**Supplementary Table 9: indicators of CKD among UK Biobank participants with African ancestry, comparing rates by number of *APOL1* variants.**

| **Genotype** | **n (total)** | **uACR >3 mg/mmol or eGFR <60 mL/min/1.73m^2^** | **uACR >3 mg/mmol** | **eGFR <60 mL/min/1.73m^2^** |
| --- | --- | --- | --- | --- |
| G0/G0 | 4,299 | 388 (9.0%) | 302 (7.0%) | 115 (2.7%) |
| G0/G1 | 2,655 | 266 (10.0%) | 209 (7.9%) | 80 (3.0%) |
| G0/G2 | 1,435 | 150 (10.5%) | 115 (8.0%) | 46 (3.2%) |
| G1/G1 | 695 | 100 (14.4%) | 81 (11.7%) | 29 (4.2%) |
| G1/G2 | 349 | 51 (14.6%) | 39 (11.2%) | 19 (5.4%) |
| G2/G2 | 161 | 18 (11.2%) | 11 (6.8%) | 12 (7.5%) |

**Supplementary Table 10: indicators of CKD among UK Biobank participants with African ancestry, comparing rates by *APOL1* genotype.**

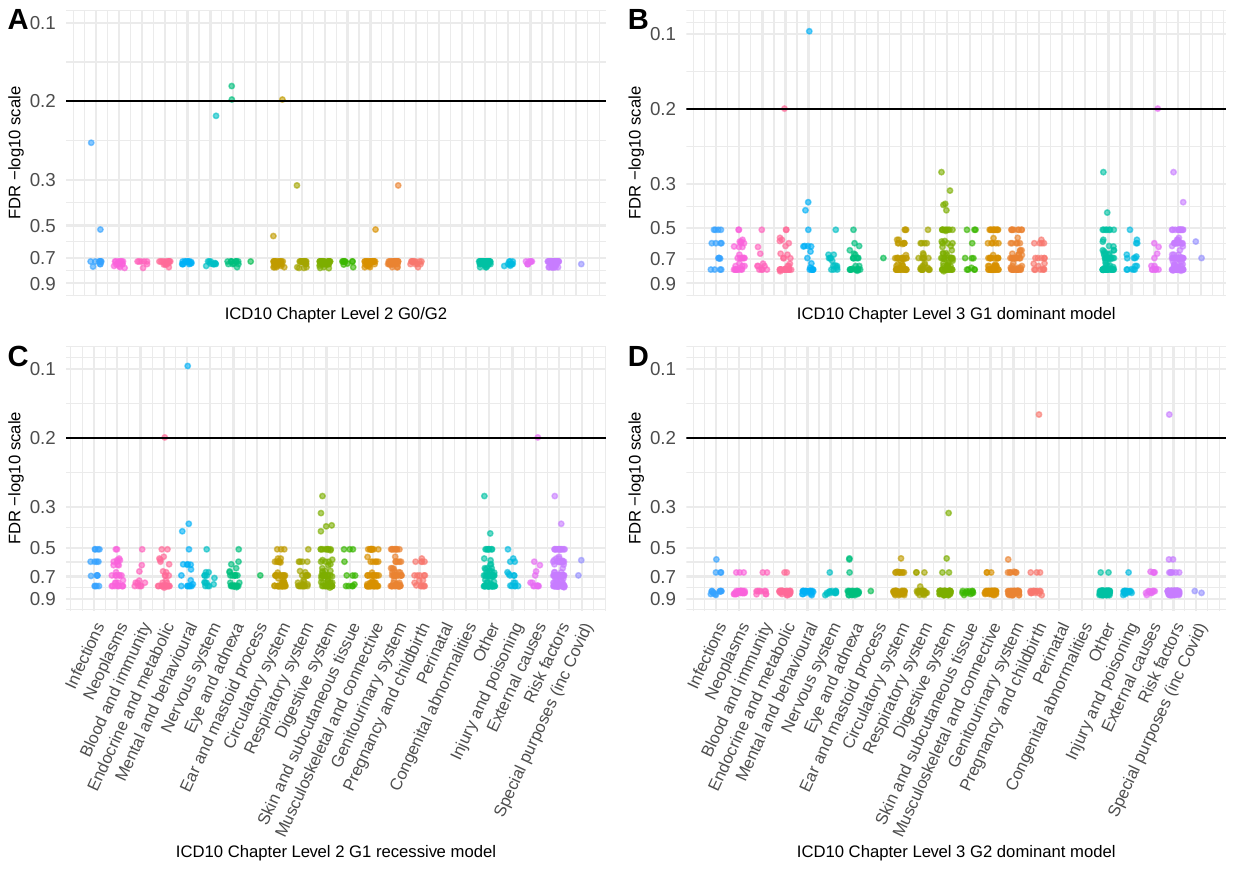

**Supplementary Figure 1: Plot of false discovery rate values showing associations between each ICD-10 code in the phenome-wide data and *APOL1* risk variant genotypes. (A) the *APOL1* G0/G2 genotype and UK Biobank Level 2 ICD-10 codes; (B) *APOL1* G1 under the dominant model (Table 5) and UK Biobank Level 3 ICD-10 codes; (C) *APOL1* G1 under the recessive model (Table 5) and UK Biobank Level 2 ICD-10 codes; (D) *APOL1* G2 under the dominant model (Table 5) and UK Biobank Level 3 ICD-10 codes. Horizontal line indicates the threshold that was used for false discovery rate (20%) for a potentially significant association.**

**Identification of end stage kidney disease**

End stage kidney disease (ESKD) as of September 2022 was defined as reaching CKD stage G5 or the requirement for kidney replacement therapy, using ICD-10 codes for hospital admission, or Office of Population Censuses and Surveys Classification of Surgical Operations and Procedures, Version 4 (OPCS4) codes for operative procedures. Participants were considered to have developed ESKD if ICD-10 codes E853, N165, N180, N185, Q601, T824, T861, Y602, Y612, Y622, Y841, Z490, Z491, Z492, Z940, Z992 , or OPCS4 codes L741, L742, L743, L744, L745, L746, L748, L749, M012, M013, M014, M015, M018, M019, M023, M084, M172, M174, M178, M179, X401, X402, X403, X404, X405, X406, X407, X408, X409, X411, X412, X418, X419, X421, X428, X429, X431 had been recorded, or if ICD-10 codes N180 or N185 appear in any position in their death record.

**Sensitivity analysis**

The ability to detect associations between the different haplotype combinations and phenotype codes using the Biobank data set for participants with African Heritage with the logistic regression model described above was estimated by simulation. Sensitivity was estimated by assigning phenotypes at random and finding the minimum odds ratio > 1 with nominal p < 0.05 observed for each haplotype combination. The same model was used as for the main analysis. Firth’s bias-reduced logistic regression was used to test the association of each phenotype with the six *APOL1* haplotype combinations. Covariates were age, sex, body mass index, Townsend deprivation index, hypertension, diabetes and the first 10 UK Biobank principal components. Given the fixed sample size, the main factors determining power in this analysis are the numbers of participants with each phenotype code and the frequency of the haplotype combinations. The deciles of the counts of phenotype codes were obtained and for each decile 1000 replicate analyses were conducted with phenotypes assigned at random to participants to obtain a range of odds ratios and p values. For each decile and haplotype combination the minimum observed odds ratio with nominal p < 0.05 was taken as an estimate of the sensitivity of the model to detect an association with that haplotype combination and that number of affected participants.

The counts of participants with each phenotype were obtained and for each decile of the counts distribution an estimate was made of the minimum odds ratio > 1 with nominal p < 0.05 that could be obtained with the model and the number of participants with each haplotype combination (Supplementary Figure 2). As expected, the minimum detectable odds ratio was inversely related to the number of participants with each haplotype combination. The G1/G2 combination which had the most associations with phenotype after the FDR correction had the second-highest minimum detectable odds ratio, indicating that the excess of associations with this haplotype combination was not due to a relatively high power to detect associations with participants with this combination. Conversely G2/G2 had a much lower power than other haplotype combinations and it is possible that associations with this haplotype combination have been underestimated due to lack of power.

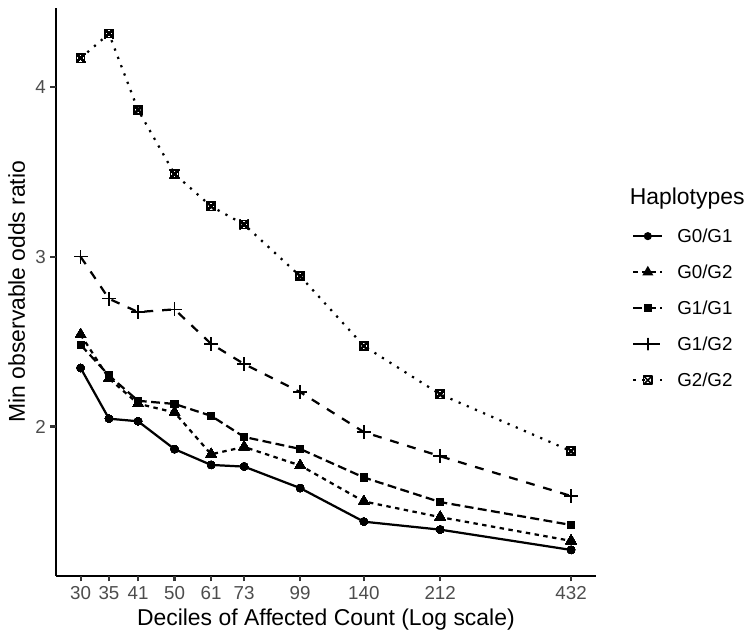

**Supplementary Figure 2. Minimum odds ratios for each haplotype combination and decile of affected counts with p < 0.05. Odds ratios and p values were generated by applying the phenome wide regression model to a dummy phenotype with ‘Count Affected’ numbers of participants being randomly assigned as cases. The affected count represents the deciles of the distribution of numbers affected.**
